# Erectile dysfunction in young men in Nairobi County, Kenya: A community-based study

**DOI:** 10.64898/2026.01.15.26344240

**Authors:** Onyango Chrispine Oluoch, Benjamin M. Ngugi, George Makalliwa

## Abstract

Erectile dysfunction is commonly perceived as a condition of older men, yet increasing evidence suggests that it also affects younger populations. Data on erectile dysfunction among young men in sub-Saharan Africa remain limited, particularly at the community level. This study assessed the prevalence of erectile dysfunction among young men in Nairobi County and examined associated sociodemographic, lifestyle, and health-related factors. A community-based cross-sectional survey was conducted among 355 sexually active men aged 18 to 35 years recruited from public service centres across Nairobi. Erectile function was assessed using a validated questionnaire, alongside measures of mental health, physical activity, medical history, and use of sexual enhancement products. Statistical analysis was used to estimate prevalence and identify factors associated with erectile dysfunction. The mean age of participants was 27.4 years. Erectile dysfunction was identified in 10.4 per cent of respondents. Symptoms of depression or anxiety were strongly associated with erectile dysfunction, as was lack of regular physical exercise. Increasing age within the study range was also associated with higher odds of erectile dysfunction. Hypertension showed a strong but inconsistent association, reflecting its low prevalence in this young population. A small but significant percentage of participants indicated the non-prescription use of sexual enhancers or erectile dysfunction medications, predominantly among those exhibiting symptoms. About one in ten young men in Nairobi suffer from erectile dysfunction, which is closely associated with psychological distress and physical inactivity. These findings highlight the necessity of incorporating sexual health, mental health screenings, and lifestyle interventions into health services for young men to address unsafe self-medication practices.

## Introduction

Erectile dysfunction (ED) is defined as the persistent inability to achieve or maintain an erection sufficient for satisfactory sexual performance (1). Although ED is often viewed as a condition in older men, it is increasingly recognized in younger men. Global prevalence estimates vary substantially, ranging from about 3 percent in some community samples to as high as 76 percent in high-risk populations, with a 2019 review estimating an overall worldwide prevalence of roughly 26 percent (2). Earlier projections suggested that the global burden of erectile dysfunction would more than double, rising from approximately 150 million men in 1995 to about 320 million by 2025, largely due to population ageing and increases in lifestyle-related risk factors, including physical inactivity (3).

Contrary to traditional belief that ED is solely an issue for older men, recent data indicate a worrying increase in ED among younger males. For example, one urology clinic in Florence, Italy observed that the proportion of men under 40 among their new ED patients tripled over a five-year period (rising from 5% in 2010 to over 15% in 2015) (4). Similarly, a large international survey reported that 8% of men aged 20–29 and 11% of men aged 30–39 had some degree of ED (5). In a U.S. cohort of young adult men, 14.2% reported ED (including mild cases) (6). These findings underscore that ED in young men, while historically understudied, is a real and common condition, not an isolation. The likely worldwide increase in ED prevalence will include many young men, highlighting the importance of understanding ED in this age group.

Multiple determinants may underlie erectile dysfunction in younger men. Historically, erectile dysfunction before age 40 was often framed primarily as psychogenic, attributed to stress, anxiety, or relationship factors, a perspective that could lead to under evaluation for organic and cardiometabolic contributors. Contemporary reviews emphasize that erectile dysfunction in young men is frequently multifactorial and warrants a systematic clinical assessment rather than presumptive reassurance (6, 7). Evidence indicates that a substantial proportion of erectile dysfunction in young men has an identifiable organic aetiology (8) reported that up to 86% of ED in men under 40 can be linked to identifiable organic causes such as vascular, neurological, or hormonal abnormalities. Metabolic and cardiovascular risk factors (e.g., hypertension, obesity, dyslipidemia) are surprisingly relevant even in this young age group, and ED in a young man can be an early warning sign of underlying cardiovascular disease (9). Mental health remains a critical component: depression and anxiety are well-known independent risk factors for sexual dysfunction. These psychological conditions can both cause and result from ED, creating a vicious cycle of performance anxiety and lowered self-esteem (10). Additionally, lifestyle behaviors such as physical inactivity, poor diet, smoking, alcohol and illicit drug use have been implicated in the pathogenesis of ED in youth by contributing to endothelial dysfunction and poor penile vascular health (11).

In Nairobi, Kenya’s capital and a major urban center, anecdotal reports from health facilities have noted an increase in young men seeking help for ED. Despite this, there has been no prior community-based study quantifying the prevalence of ED among young men in Nairobi or examining its risk factors. Past research in East Africa has focused on ED in specific high-risk groups, for instance, men with hypertension, diabetes or mental illness (who indeed have high ED rates) (12, 13). One Kenyan hospital-based study reported a 46% ED prevalence among hypertensive male patients (12). Another study in Ethiopia found ED in 94.5% of men with hypertension (14). These studies underline the impact of chronic conditions on sexual function but do not address the general young population. Furthermore, unique socio-cultural factors in Nairobi (a fast-paced urban environment) may influence sexual health differently than rural settings. A study among nomadic pastoralist men in northern Kenya documented ED correlates in that population, but their lifestyle and stressors differ greatly from urban youth (15).

Non-prescription use of sexual enhancement medicines is an emerging public health concern among young men. Sildenafil and other phosphodiesterase type 5 inhibitors, developed for clinically diagnosed erectile dysfunction, are increasingly used recreationally to enhance perceived sexual performance in men without a medical indication, often obtained without a prescription (16). A cross-sectional study among undergraduate men in Ethiopia documented phosphodiesterase type 5 inhibitor use in this population and found that use was associated with other substance use and higher-risk sexual behaviours (17). A recent systematic review similarly concludes that recreational use of oral phosphodiesterase type 5 inhibitors occurs in young, ostensibly healthy men and may be linked to maladaptive sexual expectations, potential psychological reliance, and unsafe sexual practices (18). Similarly, a survey at a Kenyan university found ∼10% of male students (mean age ∼22) had used sex-enhancing drugs or supplements (19). Such usage can mask underlying ED or even contribute to it (for example, through loss of confidence in unassisted sexual performance). The **ready availability of over-the-counter “sex enhancers”** in Nairobi, from herbal concoctions to illicitly obtained ED pills, poses public health challenges. Men may forego proper medical evaluation, and there is potential for harmful drug interactions and side effects when these substances are used without oversight.

In light of these issues, there is a clear need for data on how prevalent ED is among young men in Nairobi and what factors are associated with it. Such information can guide public health interventions, clinical screening, and education campaigns tailored to the youth. It can also inform policy, for instance, Kenya’s National Reproductive Health policy currently does not explicitly address male sexual dysfunction in young men. Filling this gap is important because untreated ED in young men can have far-reaching consequences on self-esteem, mental health, relationship stability, and quality of life. Additionally, since ED can be a harbinger of other non-communicable diseases (like cardiovascular disease or diabetes), understanding its epidemiology in young adults could enable earlier preventive healthcare.

### Study Aim

This study aimed to assess the prevalence of erectile dysfunction among sexually active men aged 18–35 in Nairobi County, Kenya, and to identify the associated sociodemographic, lifestyle, and health-related factors. We also examined the extent of non-medical use of ED medications and sexual enhancement supplements in this population. By focusing on community-dwelling young men outside of clinical patient groups, our study provides data that can help normalize conversations about ED in younger men and integrate sexual health into broader health promotion for youth.

## Materials and Methods

### Study design and setting

We conducted a community-based cross-sectional analytical study in Nairobi County, Kenya. Data collection took place in June 2025 using a one-time survey and health assessment to estimate the prevalence of erectile dysfunction and examine associations with relevant exposures among men aged 18 to 35 years. The study was observational, with no intervention administered. Participants were recruited from five Huduma Centres: GPO, City Square, Eastleigh, Kibra, and Makadara. Huduma Centres were selected because they draw large, diverse walk-in populations seeking administrative services rather than healthcare. This non-clinical setting reduced health-seeking selection bias and facilitated the recruitment of urban young men from diverse socioeconomic and residential backgrounds.

### Study Population

Eligible participants were men aged 18 to 35 years who were residents of Nairobi County for at least one year prior to the study. We included only sexually active men, defined as those who had engaged in sexual intercourse within the preceding six months. This criterion was used to ensure that ED status could be meaningfully assessed (men who had not been sexually active for an extended period were excluded to avoid misclassification of ED status due to lack of recent sexual attempts). Men were excluded if they did not meet the age range, had not lived in Nairobi for at least one year (to ensure acclimatization with the urban environment), or were unwilling/unable to provide informed consent. By focusing on the 18–35 age range, we specifically targeted the youth and young adults, aligning with the study’s objectives of exploring ED in younger men.

### Sample Strategy

A base sample size of n = **323** was determined using the single population proportion formula (Z = 1.96, d = 0.05, p = 30%) based on the estimated prevalence of erectile dysfunction among young men reported in a review by (6). Allowing for a 10% non-response rate, the final sample size was **355** participants.

Systematic sampling was used to enroll every **nth** eligible male visitor at each centre, with the sampling interval determined by the daily client flow. The goal was to evenly enroll participants across the five centers proportional to their typical daily client volume. Men who expressed interest were taken to a private area within the Huduma Centre for eligibility screening. If eligible, they were informed about the study and asked to provide written informed consent. Recruitment continued until the target sample (**N=355**) was reached. No monetary incentive was offered for participation.

### Data Collection Tools and Procedures

Data were collected using a structured, self-administered questionnaire available in English and Swahili. Participants completed the questionnaire in a private setting, with research assistants available to clarify questions. The questionnaire captured the following:

#### Sociodemographic characteristics

We collected age in years, marital status, highest education level, and employment status.

#### Lifestyle factors

Participants reported cigarette smoking, cannabis use, alcohol use, and recreational drug use history. Regular exercise was defined as moderate to vigorous physical activity at least three times per week and was recorded as yes or no.

#### Sexual behaviour and enhanced substance use

Participants reported any non-prescription use of sexual enhancers, including herbal or over-the-counter products marketed to improve erections or sexual stamina. They also reported any non-prescription use of erectile dysfunction medications, including sildenafil, tadalafil, and similar drugs. Both variables were coded as yes or no.

#### Medical history

Participants reported a history of hypertension and diabetes mellitus, recorded as yes or no for each. We also asked about prior significant physical injuries involving the pelvis, genital region, or spine, which were recorded as yes or no.

#### Psychological health

Depressive symptoms were assessed using the Patient Health Questionnaire 9, with scores of 10 or higher indicating depression. Anxiety symptoms were assessed using the Generalized Anxiety Disorder 7, with scores of 10 or higher indicating anxiety. Because depression and anxiety were strongly correlated and commonly co-occurred, we created a composite anxiety and depression variable, coded ’yes’ if either threshold was met and ’no’ otherwise.

Life satisfaction was assessed using the Life Satisfaction Questionnaire 8. Participants reporting dissatisfaction were coded as ‘no’, and those generally satisfied were coded as ‘yes’.

#### Erectile function assessment

Erectile function was assessed using the International Index of Erectile Function-5. Total scores range from 5 to 25, with higher scores indicating better erectile function. There were five levels of severity: no erectile dysfunction (25), mild (22 to 24), mild to moderate (17 to 21), moderate (12 to 16), and severe (11 or below). For prevalence and regression analyses, erectile dysfunction was defined as a score of 21 or below, and no erectile dysfunction as 22 to 25.

#### Anthropometric measurements

Weight and height were measured using a calibrated digital scale with an integrated stadiometer, with participants barefoot and wearing light clothing. The body mass index was calculated as weight in kilograms divided by height in meters squared and categorized using World Health Organization criteria as underweight, normal, overweight, or obese. Overweight and obese were combined for analysis, yielding underweight, normal weight, and overweight/obese. No blood samples or biomedical markers were collected.

#### Pre-test and data quality assurance

A pretest was conducted among 18 men aged 18 to 35 years at the GPO Huduma Centre, and these data were excluded from the main analysis. The pretest informed refinement of question wording and skip logic. Research assistants checked questionnaires for completeness at the point of collection and prompted participants to complete missing items to minimize missing data. Data were entered into a secure database and cleaned prior to analysis.

### Statistical Analysis

All completed questionnaires were checked for completeness, cleaned, coded, and entered into a secure database. The outcome variable, erectile dysfunction (ED), was coded as binary (1 = ED present, 0 = ED absent). All data were analyzed using Stata version 15.1 (StataCorp, College Station, TX). Descriptive statistics were generated to characterize the sample. We summarized continuous variables like age by mean and standard deviation and categorical variables by frequencies and percentages. The prevalence of ED was calculated as the proportion of the sample classified as having ED (IIEF-5 ≤21), with a 95% confidence interval. We also computed the prevalence (proportion) of other key variables such as depressive symptoms, anxiety, low life satisfaction, hypertension, etc., for context.

For bivariate analysis, we examined the association between each independent variable and the outcome (ED vs. no ED). We primarily used cross-tabulations and Pearson’s chi-square tests for categorical predictors. For example, we cross-tabulated ED status by categories of marital status, education level, BMI category, exercise (yes/no), etc., and obtained chi-square statistics and p-values to assess significance. These results are presented as comparisons of ED prevalence between groups. In addition, we calculated unadjusted odds ratios (OR) for having ED for each independent variable using simple logistic regression. For continuous predictors like age, univariate logistic regression was performed to estimate the OR per unit increase.

Variables that showed a statistically significant association with ED in bivariate analysis (using a threshold of p < 0.05) were selected for inclusion in a multivariable model. Given the relatively low number of ED cases (n=37), we limited the number of covariates in any single multivariable model to avoid overfitting. We combined conceptually related variables (i.e., depression and anxiety into one “anxiety/depression” variable) to reduce multicollinearity.

We ran a multivariate logistic regression using the enter method (simultaneous inclusion) to estimate adjusted odds ratios (aOR) for the predictors of ED. The final model included age, regular exercise, presence of anxiety/depression, and life satisfaction as covariates (**n=355**, since none of these had missing data). In a secondary model, we also included hypertension, but this analysis was limited to **n=235** due to missing data for hypertension (only **235** participants answered the hypertension question, because some didn’t know their blood pressure status). We thus present multiple models for comparison: one with the full sample excluding hypertension (and anxiety combined with depression) and one with the reduced sample including hypertension. All logistic models were evaluated for goodness of fit using the Hosmer-Lemeshow test, which showed no evidence of poor fit (**p greater than 0.1**). Multicollinearity was assessed using variance inflation factors, all of which were less than **2**, indicating no concerning collinearity. Model discrimination was moderate, with an area under the receiver operating characteristic curve of about **0.80**. Interaction terms, such as between exercise and depression, were examined, but none were statistically significant. Logistic regression results are presented as odds ratios with corresponding 95% confidence intervals and p-values. All tests were two-tailed, with statistical significance set at a value of 0.05.

For the third objective, the prevalence of non-prescription use of sexual enhancers and erectile dysfunction medications was calculated as simple proportions of the total sample. These outcomes were also cross-tabulated by erectile dysfunction status to explore patterns of use, although low numbers required cautious interpretation.

Results are presented in both table and narrative formats.

### Ethical Considerations

This study was approved by the KEMRI Scientific and Ethics Review Unit (SERU) (approval number **SERU/CMHS/2025/04**). A research permit was obtained from the National Commission for Science, Technology and Innovation (**NACOSTI**) prior to data collection. We also received authorization from the Huduma Kenya Secretariat and the management of each Huduma Centre involved, permitting us to recruit participants on the premises.

All participants provided written informed consent after receiving a detailed explanation of the study objectives and procedures. Participation was entirely voluntary. Participants were assured of confidentiality and the right to withdraw from the study at any point without any consequences. To protect privacy, data were collected in private rooms at the Huduma Centres, and questionnaires were self-administered (with assistance as needed) to minimize any embarrassment on sensitive questions. We employed unique study ID codes instead of names; no personally identifying information was recorded on the questionnaires.

All electronic data were stored on a password-protected computer accessible only to the principal investigator. Regular encrypted backups were made to secure cloud storage with two-factor authentication. Hard copy consent forms and any paper records were stored in a locked cabinet in the researcher’s office. Those who were identified as having severe depressive symptoms or any possible serious issue were gently advised to seek professional help, and a referral list of local health facilities (including mental health and urology services) was provided.

No physical risks were anticipated in this survey.

## Results

### Participant Characteristics

A total of 355 men, ages 18–35, were recruited from five Huduma centres in Nairobi County. Mean age was 27.4 years (SD 5.1). Most were single (54.4%), had tertiary education (62.0%), and were employed (61.1%). The distribution of body mass index was 9.9% underweight, 63.4% normal weight, and 26.8% overweight or obese. Hypertension and diabetes mellitus were uncommon at 1.7% and 0.3%, respectively. Prior significant perineal or genital trauma occurred in 11.8% and showed no association with erectile dysfunction. Regular exercise at least three times per week was reported by 67.3%, while 32.7% did not exercise regularly, and 31.0% indicated recreational drug use. Depressive symptoms were present in 24.2% and anxiety symptoms in 18.9%; these were moderately correlated (r = 0.60, p < 0.001), hence the use of a composite "anxiety/depression" variable in subsequent analyses. Life satisfaction was high overall, with 9.3% dissatisfied. Sociodemographic characteristics are presented in **Table 1**.

**Table 1.** Sociodemographic characteristics of the study participants (N = 355)

| Variable | Category | n (%) |
| --- | --- | --- |
| Age, years | Mean (SD) | 27.4 (5.1) |
| Marital status | Single | 193 (54.4) |
|  | Married or cohabiting | 162 (45.6) |
| Education level | Never attended school | 2 (0.6) |
|  | Primary | 15 (4.2) |
|  | Secondary | 118 (33.2) |
|  | Tertiary | 220 (62.0) |
| Employment status | Employed | 217 (61.1) |
|  | Not employed | 138 (38.9) |
| Body mass index category | Underweight | 35 (9.9) |
|  | Normal weight | 225 (63.4) |
|  | Overweight or obese | 95 (26.8) |

**Abbreviations:** SD, standard deviation.

### Prevalence of Erectile Dysfunction

Out of 355 participants, **37 men were classified as having erectile dysfunction**, corresponding to an overall ED prevalence of **10.4%** (95% confidence interval 7.4%–14.1%). This prevalence encompasses all degrees of ED severity. Breaking it down by severity via IIEF-5 scores: 18 men (5.1%) had mild ED, 8 men (2.3%) had mild-to-moderate ED, 6 men (1.7%) had moderate ED, and 5 men (1.4%) had severe ED. The remaining 318 men (89.6%) had no ED (IIEF-5 scores in the normal range).

To our knowledge, this is the first community-derived prevalence estimate for ED in young Kenyan men. The 10.4% figure suggests that roughly one in ten men under 35 in Nairobi experiences at least occasional or mild erectile dysfunction.

### Factors Associated with ED: Bivariate Analysis

Age showed a clear positive association with ED, with a 13% increase in odds per year (OR 1.13, 95% CI 1.05 - 1.22, p = 0.001). Hypertension was associated with markedly higher odds of ED (OR 8.54, 95% CI 1.63 - 44.71, p = 0.011), although estimates were imprecise due to few hypertensive participants. Diabetes mellitus was excluded because it perfectly predicted ED, as the only participant with diabetes reported it. Psychological distress was strongly associated with ED, with approximately fivefold higher odds among men screening positive for anxiety or depression (OR 5.05, 95% CI 2.48 - 10.30, p < 0.001). Regular exercise was protective, with 68% lower odds among men who exercised (OR 0.32, 95% CI 0.16 - 0.65, p = 0.001). A greater sense of life satisfaction correlated with reduced odds of erectile dysfunction (OR 0.38, 95% CI 0.15 - 0.95, p = 0.039); however, this correlation diminished when controlling for anxiety or depression, indicating potential mediation by mental health factors. The results of bivariate analysis are presented in **Table 2**.

**Table 2.** Bivariate associations between participant characteristics and erectile dysfunction.

| Variable | Odds Ratio (OR) | 95% Confidence Interval | p-value |
| --- | --- | --- | --- |
| Age (years) | 1.13 | 1.05 – 1.22 | 0.001 |
| BMI category | 1.42 | 0.98 – 2.05 | 0.061 |
| Education level | 1.46 | 0.77 – 2.75 | 0.246 |
| Marital status | 1.46 | 0.74 – 2.89 | 0.279 |
| Employment status | 1.82 | 0.85 – 3.89 | 0.123 |
| Hypertension (HTN) | 8.54 | 1.63 – 44.71 | 0.011 |
| Diabetes mellitus | — | — | — |
| History of physical injury | 1.19 | 0.44 – 3.24 | 0.738 |
| Regular physical exercise | 0.32 | 0.16 – 0.65 | 0.001 |
| Recreational drug use | 1.82 | 0.91 – 3.63 | 0.093 |
| Anxiety/Depression | 5.05 | 2.48 – 10.30 | <0.001 |
| Life satisfaction | 0.38 | 0.15 – 0.95 | 0.039 |

**Notes:** Diabetes mellitus perfectly predicted erectile dysfunction because only one participant reported diabetes and had erectile dysfunction, therefore an odds ratio could not be estimated.

### Multivariate Predictors of Erectile Dysfunction

We fitted three multivariable logistic regression models to identify independent predictors of ED while controlling for confounding. Some participants reported that they did not know their hypertension status; these responses were coded as missing, leaving hypertension data for **235** participants.

**Model 1** included age, hypertension, regular exercise, anxiety/depression, and life satisfaction (n = 235). Age was independently associated with ED (aOR 1.13, 95% CI 1.02 - 1.25, p = 0.018). Regular exercise was protective (aOR 0.37, 95% CI 0.15 - 0.92, p = 0.031), while anxiety/depression strongly predicted ED (aOR 5.67, 95% CI 2.23 - 14.45, p < 0.001). Hypertension and life satisfaction were not significant.

**Model 2** excluded anxiety/depression to examine the effect of hypertension (n = 235). Age remained significant (aOR 1.13, 95% CI 1.02 - 1.24, p = 0.016), exercise remained protective (aOR 0.39, 95% CI 0.17 - 0.92, p = 0.031), and hypertension became significant (aOR 8.07, 95% CI 1.37 - 47.67, p = 0.021). Life satisfaction was not associated.

**Model 3** excluded hypertension to retain the full sample and served as the primary model (n = **355**). Age remained associated with ED (aOR 1.12, 95% CI 1.03 - 1.21, p = 0.005), exercise remained protective (aOR 0.37, 95% CI 0.18 - 0.77, p = 0.008), and anxiety/depression remained the strongest predictor (aOR 5.13, 95% CI 2.36 - 11.18, p < 0.001). Life satisfaction was not significant.

Overall, age, exercise, and anxiety/depression showed consistent independent associations with ED across models, while life satisfaction did not. Results are presented in **Table 3**.

**Table 3.** Multivariate logistic regression models for predictors of erectile dysfunction among young men in Nairobi County.

| Variable | Model 1 – aOR (95% CI) | Model 2 – aOR (95% CI) | Model 3 – aOR (95% CI) |
| --- | --- | --- | --- |
| Age (per year) | 1.13 (1.02–1.25)<br>$p = 0.018$ | 1.13 (1.02–1.24)<br>$p = 0.016$ | 1.12 (1.03–1.21)<br>$p = 0.005$ |
| Hypertension | 5.50 (0.72–42.01)<br>$p = 0.100$ | 8.07 (1.37–47.67)<br>$p = 0.021$ | — (*excluded*) |
| Regular exercise | 0.37 (0.15–0.92)<br>$p = 0.031$ | 0.39 (0.17–0.92)<br>$p = 0.031$ | 0.37 (0.18–0.77)<br>$p = 0.008$ |
| Anxiety/ Depression | 5.67 (2.23–14.45)<br>$p < 0.001$ | — (*excluded*) | 5.13 (2.36–11.18),<br>$p < 0.001$ |
| Life satisfaction | 0.92 (0.25–3.43)<br>$p = 0.899$ | 0.47 (0.14–1.62)<br>$p = 0.234$ | 0.78 (0.28–2.20),<br>$p = 0.644$ |
| **Model stats:** | N = 235; LR $\chi^2(5) = 33.00$, $p < 0.001$;<br>Pseudo R<sup>2</sup> = 0.197 | N = 235; LR $\chi^2(4) = 18.98$, $p = 0.001$;<br>Pseudo R<sup>2</sup> = 0.113 | N = 355; LR $\chi^2(4) = 39.18$, $p < 0.001$;<br>Pseudo R<sup>2</sup> = 0.165 |

**Abbreviations:** aOR, adjusted odds ratio; CI, confidence interval.

The study also assessed the use of non-prescribed erectile dysfunction-related products. Twenty participants (5.6%) reported ever using sexual enhancement supplements or herbs, and 17 (4.8%) reported using prescription erectile dysfunction medicines without a prescription. The majority of users were men with erectile dysfunction; 16 of the 17 non-prescribed medicine users were classified as having erectile dysfunction on the IIEF-5, which suggests that they were using the medicine to treat their symptoms.

In summary, erectile dysfunction affected about one in ten young men in Nairobi County. Independent predictors were increasing age, lack of regular exercise, and anxiety/depression. Life satisfaction was not independently associated, and hypertension showed inconsistent associations across models.

## Discussion

This community-based study provides novel epidemiologic evidence on erectile dysfunction (ED) among men aged 18–35 years in an urban African setting. Increasing age within this young cohort was associated with higher odds of ED, as were modifiable factors including lack of regular physical exercise and the presence of anxiety or depression.

In Nairobi County, ED prevalence was 10.4%, broadly consistent with international community estimates for younger men. The multinational MALES study reported ED rates of 8% among men aged 20–29 years and 11% among those aged 30–39 years, while a population-based survey in São Paulo, Brazil, found a prevalence of 7.3% among men aged 20–29 years (20, 21). Our observed ED prevalence lies at the lower end of estimates reported in recent studies of young male populations; for example, a U.S. cohort of men younger than 40 years reported a prevalence of 14.2% (22). Other reports have noted rates as high as 30–35% in young men, though such higher estimates often depend on specific populations (23). Thus, our estimate that roughly one in ten young men in Nairobi has at least mild ED appears generally consistent with global data, and it underscores that ED is not uncommon even in young adulthood. Notably, epidemiologic data specific to sub-Saharan Africa on young men’s sexual health are scarce. Most African studies have focused on older age groups or special clinical populations, which show much higher ED rates (often 30–60% or more) (12, 14, 24, 25). For instance, community and clinic-based studies report ED prevalence of about 29.7% in Tanzania and 58–59% in Nigeria (26, 27). To our knowledge, this is the first study to quantify ED prevalence among Kenyan men under 35 years outside a clinical setting.

Within this narrow age band, ED increased progressively with advancing age, consistent with prior evidence that ED risk rises continuously across the adult life course (28, 29). This pattern may reflect accumulated exposures in the third decade of life, including early cardiometabolic changes, sustained psychosocial stressors, and prolonged adverse health behaviours.

Physical activity was strongly protective in our analysis, corroborating evidence that sedentary lifestyles contribute to sexual dysfunction. Regular exercise supports erectile physiology through improved vascular and endothelial function, enhanced nitric oxide bioavailability, and favorable hormonal regulation, all of which together help preserve normal erectile function (30, 31). A large Harvard prospective cohort reported roughly a 30% lower risk of incident ED among men engaging in vigorous exercise compared with their sedentary peers (32), and a Brazilian study of nearly 21,000 men found higher physical activity to be independently associated with reduced odds of ED after adjustment for comorbidities (28). These convergent findings support physical activity promotion as a practical target for preserving sexual health in young adulthood.

Psychological factors emerged as the strongest predictors of ED in this young cohort: men screening positive for anxiety and/or depression had more than fivefold higher odds of ED. This aligns with contemporary evidence that ED in younger men is frequently multidimensional, with psychogenic contributors such as performance anxiety, depressive symptoms, and stress often playing a dominant role (33). In an urban African setting where mental health stigma remains substantial, these findings underscore the likelihood that unaddressed psychosocial stressors may be expressed through sexual dysfunction and support integrating routine mental health screening into ED evaluation and management.

Interestingly, life satisfaction – a broader measure of overall well-being – was protective in unadjusted analysis but not an independent factor in multivariate models once anxiety/depression was accounted for. This suggests that general life contentment alone does not buffer against ED if specific psychological distress is present; targeted mental health factors are more proximal determinants of ED than a global happiness measure.

Hypertension in our study showed a positive association with ED in certain models (aOR ∼5–8), consistent with the known pathophysiological connection between vascular health and erectile function (34, 35). However, because hypertension was relatively uncommon in this young cohort (∼1.7% prevalence), its effect was unstable. We interpret this cautiously: a young man with hypertension likely has elevated ED risk (as seen in older populations), but our study lacked power to conclusively confirm hypertension as an independent predictor.

Similarly, diabetes mellitus was extremely rare in this cohort: only one participant reported diabetes, and he had ED, resulting in perfect prediction and preventing estimation of an odds ratio. Although based on a single case, this pattern is consistent with the broader evidence that ED can be an early marker of underlying metabolic dysfunction, including insulin resistance and diabetes (36).

Several factors were not significantly associated with ED in this young population, including BMI, education level, marital status, employment status, history of physical injury, and recreational drug use. Although recreational drug use would be expected in this age group and is recognized as an ED risk factor (37–42), the null association here may reflect low cumulative exposure and possible underreporting due to social desirability bias. Overall, these results suggest that physical inactivity and psychological distress were more salient correlates of ED than the measured sociodemographic factors or recreational drug use.

### Public Health, Clinical, and Policy Implications

These findings indicate that ED among young men warrants public health attention with clear clinical and policy relevance, particularly in Kenya and comparable settings. A prevalence of about one in ten suggests a meaningful burden that may be underestimated because of stigma, supporting inclusion of ED within youth-friendly health services and outreach. Clinically, ED in this age group should be managed within a biopsychosocial framework and prompt a structured, nonjudgmental assessment that includes mental health screening, lifestyle counselling, and targeted cardiometabolic risk evaluation, with opportunistic screening emphasized for men reporting psychological distress, physical inactivity, or elevated blood pressure. Policy priorities include strengthening youth-friendly mental health services, promoting physical activity through population-based interventions, expanding sexual health education that normalizes help-seeking, and improving regulation and pharmacovigilance for ED medicines and sexual enhancement products to reduce unsafe self-medication while ensuring access to appropriate care.

### Study Strengths

Key strengths of this study include a community-based, multisite design and the use of validated instruments (IIEF-5, PHQ-9, GAD-7, and LISAT-8), which enhance measurement standardisation. Additionally, the self-administered questionnaire approach likely mitigated interviewer-related bias by reducing direct interactions between participants and data collectors.

### Limitations and Future Directions

Limitations include the cross-sectional design, which precludes causal inference and limits temporal interpretation of ED–mental health relationships; reliance on self-report, which may underestimate prevalence; restriction to an urban sample; and lack of hormonal or vascular measures to distinguish organic from psychogenic ED.

Future research should extend to multicentric national cohorts, including rural and peri-urban populations; incorporate longitudinal designs to clarify temporality; and evaluate integrated lifestyle and mental health interventions.

## Conclusion

This community-based study indicates that erectile dysfunction affects about one in ten men aged 18 to 35 years in Nairobi and is consistently associated with physical inactivity and psychological distress. The concentration of unprescribed medication use among affected men highlights unmet, stigma-related barriers to appropriate care. Erectile dysfunction in early adulthood should be treated as a sentinel marker for broader health vulnerability and an entry point for integrated prevention-focused services that combine mental health assessment, physical activity promotion, and targeted cardiometabolic risk evaluation.

## Acknowledgments

The authors thank all the men who participated in this study for their time and openness in responding to sensitive questions. Appreciation is extended to the staff and secretariat of the Nairobi Huduma Centres for facilitating the research activities. We also extend our sincere gratitude to the field research assistants who collected the data with strict respect for the participants’ privacy.

## Author Contributions

Onyango Chrispine Oluoch (OCO) conceived and designed the study, obtained ethical approval, developed the data collection instruments, coordinated data collection, conducted the statistical analyses, and drafted the manuscript. Benjamin M. Ngugi (BMN) and George Makalliwa (GM) provided methodological guidance, contributed to the literature review, critically interpreted the findings, and reviewed and scrutinized the data analysis and manuscript. All authors approved the final version of the manuscript and accept responsibility for its content.

## Conflict of Interest Statement

The authors declare no conflicts of interest. This research was conducted independently as part of academic requirements and was not influenced by any external commercial or financial interests. There was no funding from pharmaceutical companies or entities that could present a conflict. All analyses and interpretations are those of the authors.

## Data Availability Statement

The de-identified data supporting the findings of this study are available from the corresponding author on reasonable request. Participants’ privacy and consent conditions will be upheld in determining data sharing. The study questionnaire and codebook are also available on request.

## Notes

### Competing Interest Statement

The authors have declared no competing interest.

### Funding Statement

No grants or financial support of any kind were received for this study. The research was conducted independently and was not influenced by any commercial or financial interests.

### Author Declarations

This study was approved by the Kenya Medical Research Institute Scientific and Ethics Review Unit (KEMRI SERU). A research permit was obtained from the National Commission for Science, Technology and Innovation (NACOSTI). Additional authorisation was granted by the Huduma Kenya Secretariat and the management of the participating Huduma Centres.

